# Applying the Three Delays Model to Understand Care Pathway Barriers among Low-Birth-Weight Neonates in a Kenyan Referral Hospital

**DOI:** 10.64898/2026.08.03.26359628

**Authors:** Judy Cheptoo, Morris Senghor Shisanya, Vincent K. Mukthar, Everlyne Morema

## Abstract

**Background:** Outcomes for low-birth-weight (LBW) neonates depend not only on biology but on the timeliness of the care pathway. The Three Delays Model—deciding to seek care (Delay 1), reaching the hospital (Delay 2), and receiving adequate care after arrival (Delay 3)—offers a validated lens for locating where that pathway fails. We applied the model to characterise care-pathway barriers affecting LBW neonates admitted to a Kenyan county referral hospital and to relate them to severe adverse outcomes.

**Methods:** Facility-based mixed-methods cross-sectional study of 169 LBW neonate–mother pairs admitted to the newborn unit of Kericho County Referral Hospital, complemented by nine key-informant interviews with providers. Delay indicators were derived for each of the three delays, with denominators defined explicitly. Descriptive statistics summarised each indicator; associations with severe adverse outcome were tested with the chi-square or Fisher exact test (kept descriptive, not modelled). Provider interviews were analysed thematically and coded directly to the three delays; quantitative and qualitative findings were integrated in a delay-structured joint matrix.

**Results:** A severe adverse outcome occurred in 136/169 neonates (80.5%). Pathway barriers clustered before arrival: decision-to-seek-care delay >6 h in 13.6%, a transport-access problem in 32.5%, and residence >10 km from a facility in 34.3%; nearly half (49.1%) were referred/outborn, and among referred neonates 26.5% arrived without a referral note. After arrival, care began within 30 minutes in 66.3%. Referral/outborn status was associated with higher odds of a severe outcome (crude OR 2.25, 95% CI 1.01–5.00; p = 0.043), as was essential drug/feed shortage (OR 2.26, 95% CI 1.04–4.90; p = 0.036). Paradoxically, decision delay, transport problems, and any pathway delay were each associated with a lower proportion of severe outcomes (all p < 0.01); these inverse associations most plausibly reflect confounding by indication and reverse causation—the sickest neonates were prioritised for rapid transfer and care—and should not be read as protective effects of delay. Provider narratives, coded to the three delays, described caregiver danger-sign recognition gaps, transport and referral-coordination barriers (cold, poorly documented arrivals), and first-hour stabilisation, staffing, warm-chain, supply, and monitoring constraints.

**Conclusions:** Barriers for the smallest neonates accumulate along the pre-hospital pathway, and referral status signals more than a transport category—it marks accumulated vulnerability from delayed decision-making, transport constraints, incomplete pre-referral stabilisation, and facility-response gaps. Reducing severe outcomes requires shortening specific, identifiable delays, especially strengthening referral coordination and the fragile first hour after arrival, rather than reproducing a full determinants model.

## Introduction

Low birth weight (LBW; <2,500 g) remains one of the strongest markers of neonatal vulnerability worldwide [1], and preterm and small newborns account for a disproportionate share of the roughly 2.3 million neonatal deaths that occur each year, most of them in sub-Saharan Africa and South Asia [2,3]. For these infants, the interval immediately after birth or referral is decisive: hypothermia, hypoglycaemia, sepsis, and respiratory compromise can convert a manageable biological vulnerability into a severe adverse outcome within hours [4–7]. Survival therefore depends not only on how small or preterm a neonate is, but on how quickly and reliably the care pathway responds.

The Three Delays Model, first articulated by Thaddeus and Maine to explain maternal deaths [8], locates preventable mortality within three sequential points of failure: a delay in the decision to seek care (Delay 1), a delay in reaching an appropriate facility (Delay 2), and a delay in receiving adequate care after arrival (Delay 3). Although developed for maternal health, the framework has been productively extended to newborn survival—most notably by Waiswa and colleagues in eastern Uganda [9]—and subsequently applied to neonatal deaths across sub-Saharan Africa and South Asia [10–13], because the newborn, like the mother, depends on a chain of timely decisions and actions by caregivers, transporters, and providers. Applied to LBW neonates, the model reframes the clinical question from “which factors are associated with poor outcomes” to “where along the pathway does care break down, and which break points are locally modifiable” [14].

County referral hospitals in Kenya sit at the convergence of this pathway. They receive both inborn LBW neonates and outborn neonates referred from lower-level facilities, often after variable pre-referral stabilisation and transport [15]. In such settings, biological vulnerability and health-system readiness intersect at the point of admission, and the first hour of care is frequently the moment at which trajectories are set [4,16]. Yet facility-level evidence on where the neonatal care pathway fails—and on how quantitative delay indicators relate to what providers actually experience at the bedside—remains limited for many Kenyan county referral units [17–19].

This paper applies the Three Delays Model to LBW neonates admitted to the newborn unit of Kericho County Referral Hospital (KCRH). Rather than reproducing a full determinants model, we deliberately confine the analysis to the care pathway: care-seeking and decision delays, transport and referral barriers, and first-hour triage and stabilisation. Our aim was to characterise the care-seeking, referral, transport, triage, and facility-level delays affecting LBW neonates, to relate these delays to severe adverse outcomes, and to integrate them with provider perspectives so that the most actionable delay for this setting can be identified.

## Materials and Methods

### Study design and setting

We conducted a facility-based, mixed-methods cross-sectional study at the newborn unit of Kericho County Referral Hospital, a high-volume referral facility for high-risk neonates in Kericho County, Kenya between 3^rd^ April 2026 and 7^th^ Jun 2026. The quantitative strand characterised the care pathway of admitted LBW neonates; the qualitative strand captured provider perspectives on the same pathway. The study is reported in line with STROBE for the observational quantitative component, COREQ for the qualitative component, and GRAMMS for mixed-methods integration.

### Theoretical framework

The Three Delays Model provided the organising structure for the entire study. Every variable, analysis, table, and figure was assigned a priori to one of three delays: Delay 1 (deciding to seek care), Delay 2 (reaching the hospital), and Delay 3 (receiving adequate care after arrival). This structure governs both the results and the discussion. Consistent with the model’s original intent, the delays are treated as sequential and cumulative points at which a neonate’s condition may deteriorate.

### Participants and sampling

The analytic sample for the quantitative strand comprised 169 LBW neonate–mother pairs admitted to the newborn unit during the study period, each with care-pathway and delay data. For the qualitative strand, nine healthcare workers with direct newborn-care responsibilities were purposively sampled to capture a range of cadres and experience: one paediatrician, five nurses, two clinical officers, and one newborn-unit manager, with 2–10 years of neonatal experience. Sampling continued until the provider accounts of the care pathway reached thematic saturation.

### Delay indicators and denominators

From the pathway variables collected, we derived a set of binary and categorical delay indicators, each mapped to a single delay and each with an explicitly defined denominator. Delay 1 indicators were derived from the reported interval from onset of labour/illness to the decision to seek care (delay defined as >6 h), the person who made the final care-seeking decision, and maternal involvement in that decision. Delay 2 indicators were derived from residence, distance to facility (long distance defined as >10 km), travel time (delay defined as >2 h), means of transport, self-reported transport-access problems, referral/outborn status, and referral coordination (referral note provided or not). Delay 3 indicators were derived from the time from admission to initiation of neonatal care (prompt care defined as <30 min; delayed initiation as >1 h), admission to the newborn unit, availability of skilled personnel at admission, maintenance of the warm chain, and shortage of essential neonatal drugs or feeds. Unless otherwise stated, the denominator was the full cohort (N = 169); referral coordination among referred neonates used the referred subgroup (n = 83) as its denominator. A delay-indicator codebook documenting each derivation and denominator is provided in the Supporting Information (S1).

### Quantitative analysis

Delay indicators were summarised as frequencies and percentages, and continuous or ordinal indicators (e.g., number of delay factors) as medians with interquartile range (IQR), grouped by delay stage. Associations between key delay indicators and severe adverse outcome were tested with the Pearson chi-square test, or the Fisher exact test where expected cell counts were small, and are reported with crude odds ratios (ORs) and 95% confidence intervals (CIs). In keeping with the paper’s scope, association testing was deliberately kept descriptive: we did not fit an adjusted multivariable determinants model, which is reserved for the companion determinants paper. The primary outcome, severe adverse neonatal outcome, was a composite defined a priori as the occurrence of neonatal death, or a prolonged newborn-unit stay (≥7 days), or two or more concurrent adverse outcomes among respiratory distress, sepsis, hypothermia, hypoglycaemia, low Apgar score, and developmental delay. Analyses were conducted in IBM SPSS Statistics; a two-sided p < 0.05 was considered statistically significant.

### Qualitative analysis

Key-informant interviews were audio-recorded, transcribed, and analysed thematically following Braun and Clarke. Coding was framework-guided: an initial deductive frame reflecting the three delays was combined with inductive codes emerging from the data. Two analysts coded independently, met to reconcile codes, and maintained a coding audit trail and reflexive notes. Themes were then mapped directly to Delay 1, Delay 2, or Delay 3.

### Integration

Integration followed a delay-structured joint-display approach. Within each delay, the relevant quantitative indicator(s) were placed alongside the corresponding qualitative theme and an integrated interpretation, allowing convergence, divergence, and measurement gaps to be read directly (Table 5). Where quantitative and qualitative findings pointed in opposite directions—as occurred for several delay indicators—the joint display was used to reason explicitly about confounding by indication and reverse causation rather than to force agreement.

### Ethics

The study was approved by the by Kabarak University Research Ethic Committee (KUREC) (approval number KUREC-170226) and licensed by the National Commission for Science, Technology and Innovation (NACOSTI) (License number NACOSTI/P/26/4186446), and administrative authorization from Kericho County Referral Hospital (P/26/4186446) was obtained from the hospital. Written informed consent was obtained from mothers/guardians for the neonatal record component and from providers for the interviews. Data were de-identified before analysis, and confidentiality was maintained throughout.

### Data availability

The de-identified dataset and the analysis syntax supporting the findings are available from the corresponding author on reasonable request, subject to the conditions of the ethical approval. A delay-indicator codebook is provided as Supporting Information.

## Results

### The care pathway and its outcome

All 169 neonates were low birth weight, and a severe adverse outcome occurred in 136 (80.5%). Against this high baseline burden, the pathway analysis asks where—between the home and the first hour of newborn-unit care—the barriers to timely care were concentrated. The distribution of delay indicators across the three delays is summarised in Table 1 and displayed in Figure 2; associations with the severe outcome are reported in Tables 2 and 3; the qualitative themes and the integrated joint matrix follow in Tables 4 and 5; and the adapted pathway is depicted in Figure 1.

**Figure 1.**
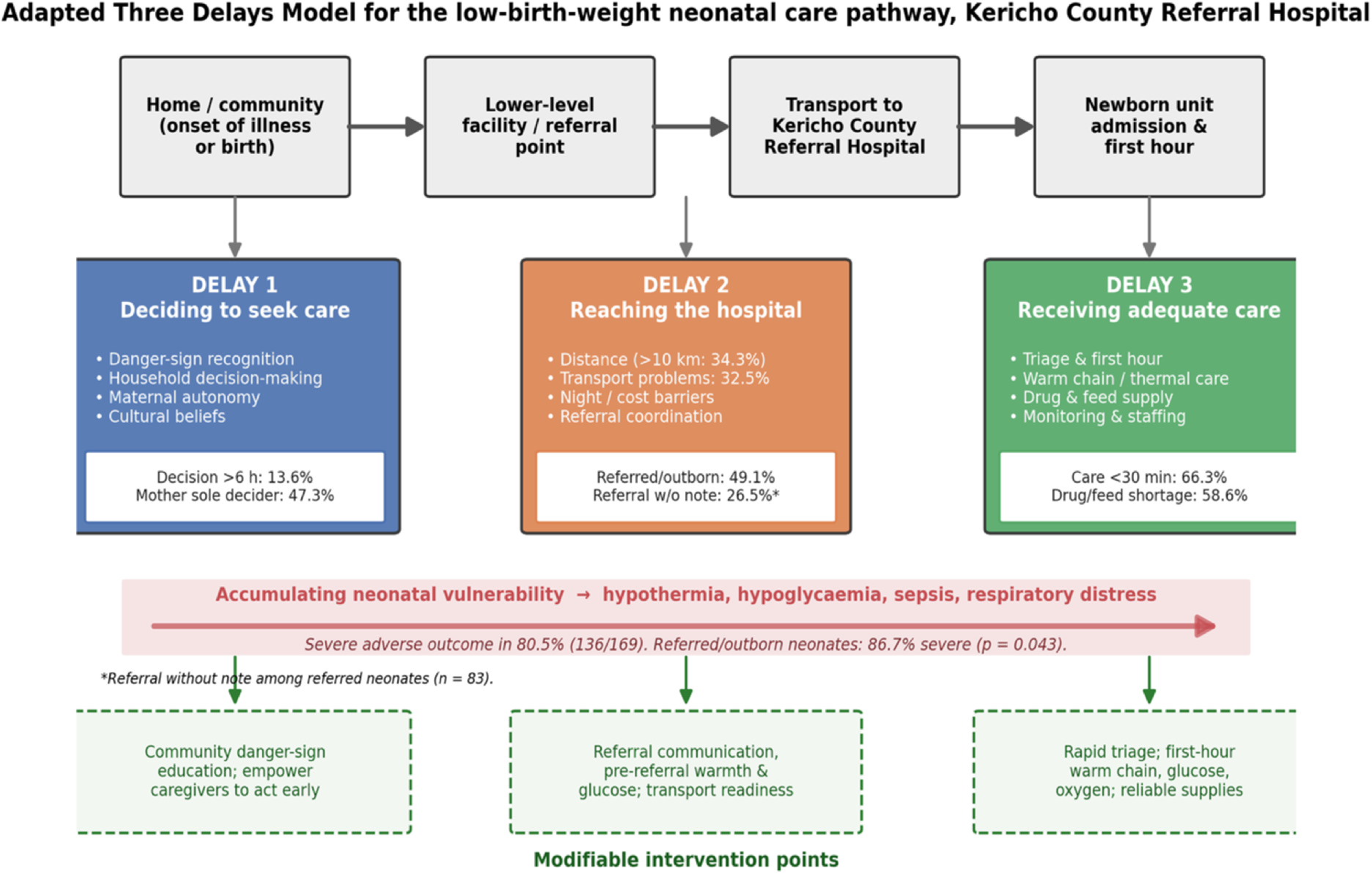
Adapted Three Delays Model for the low-birth-weight neonatal care pathway at Kericho County Referral Hospital. The pathway runs from the home/community, through a lower-level facility and transport, to newborn-unit admission and the first hour of care. At each delay, the setting-specific barriers, the accumulating neonatal vulnerability, and the modifiable intervention points are shown. Referred/outborn neonates carried a higher proportion of severe outcomes (86.7%).

**Figure 2.**
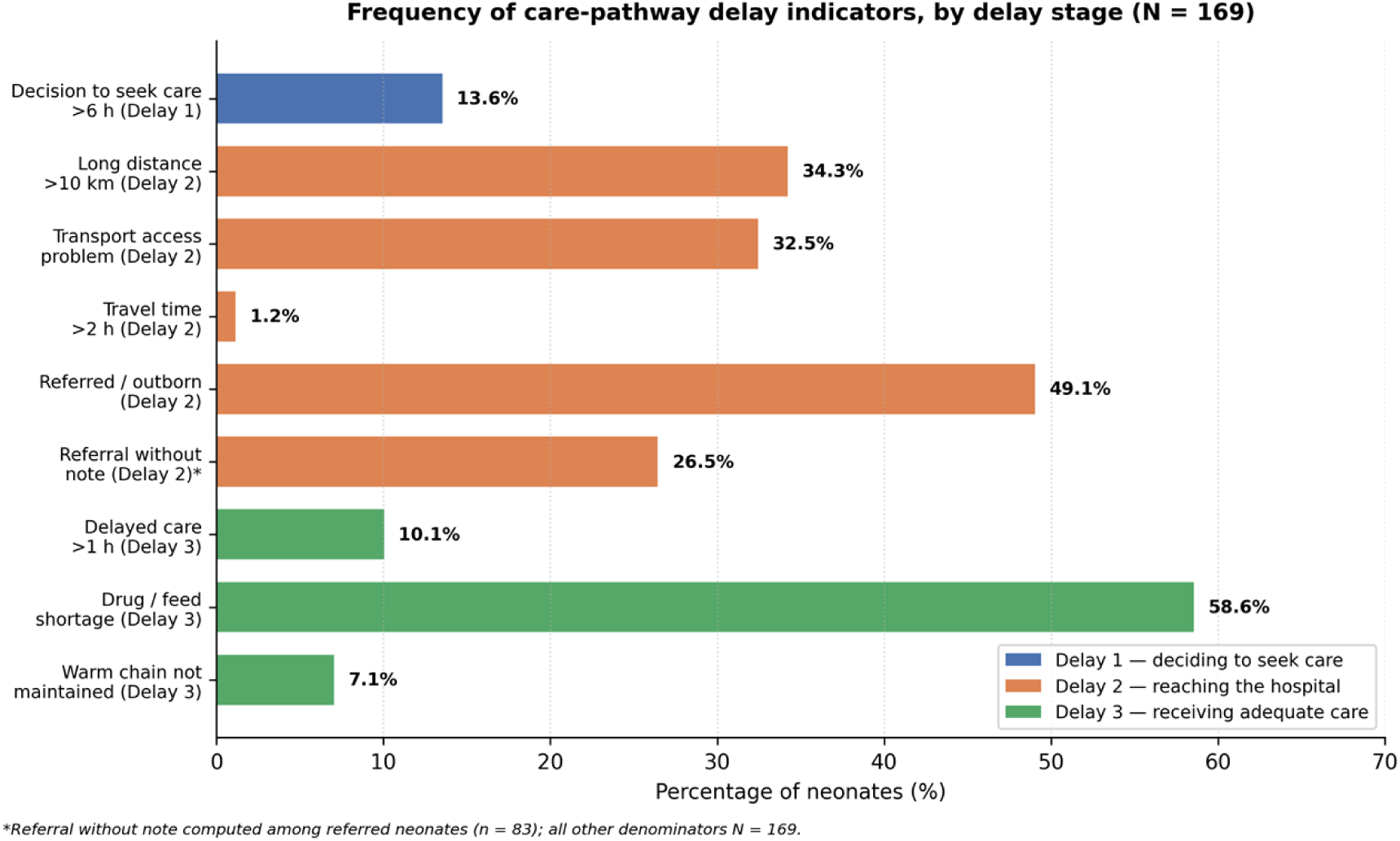
Frequency of the main care-pathway delay indicators, by delay stage (N = 169). Delay 2 (reaching the hospital) accounts for the most prevalent pathway barriers—long distance, transport problems, and referral—while the facility-level delay (>1 h to care) was comparatively uncommon.

**Table 1.** Care-seeking and delay characteristics organised by delay stage (N = 169)

| Delay indicator | n (%) or median (IQR) | Denominator |
| --- | --- | --- |
| ***Delay 1 — Deciding to seek care*** |  |  |
| Onset-to-decision: immediately | 80 (47.3%) | 169 |
| Onset-to-decision: within 6 h | 66 (39.1%) | 169 |
| Onset-to-decision: >6 h (decision delay) | 23 (13.6%) | 169 |
| Final decision-maker: mother alone | 80 (47.3%) | 169 |
| Final decision-maker: joint | 48 (28.4%) | 169 |
| Final decision-maker: partner | 26 (15.4%) | 169 |
| Final decision-maker: other | 15 (8.9%) | 169 |
| Mother involved in care-seeking decision | 128 (75.7%) | 169 |
| ***Delay 2 — Reaching the hospital*** |  |  |
| Rural residence | 99 (58.6%) | 169 |
| Distance to facility: <5 km | 60 (35.5%) | 169 |
| Distance to facility: 5–10 km | 51 (30.2%) | 169 |
| Distance to facility: >10 km (long distance) | 58 (34.3%) | 169 |
| Travel time >2 h (reaching delay) | 2 (1.2%) | 169 |
| Transport: motorbike | 67 (39.6%) | 169 |
| Transport: private car | 63 (37.3%) | 169 |
| Transport: public transport | 32 (18.9%) | 169 |
| Transport: on foot | 4 (2.4%) | 169 |
| Transport: ambulance | 3 (1.8%) | 169 |
| Transport-access problem reported | 55 (32.5%) | 169 |
| Referred / outborn | 83 (49.1%) | 169 |
| Referred with referral note | 61 (36.1%) | 169 |
| Referred without referral note | 22 (13.0%) | 169 |
| Referral without note (among referred) | 22 (26.5%) | 83 |
| ***Delay 3 — Receiving adequate care after arrival*** |  |  |
| Admission-to-care: <30 min (prompt care) | 112 (66.3%) | 169 |
| Admission-to-care: 30–60 min | 40 (23.7%) | 169 |
| Admission-to-care: >1 h (delayed care) | 17 (10.1%) | 169 |
| Admitted to newborn unit (NBU/NICU) | 160 (94.7%) | 169 |
| Skilled personnel available at admission | 160 (94.7%) | 169 |
| Warm chain maintained | 157 (92.9%) | 169 |
| Essential drug/feed shortage | 99 (58.6%) | 169 |
| ***Cross-cutting*** |  |  |
| Any pathway delay present | 72 (42.6%) | 169 |
| Number of delay factors, median (IQR) | 0 (0–1) | 169 |

*Note.* Values are n (%) unless otherwise indicated. Delay 1 (deciding to seek care), Delay 2 (reaching care), and Delay 3 (receiving care) follow the Three Delays Model. Decision delay = onset-to-decision >6 h; long distance = >10 km; reaching delay = travel time >2 h; prompt care = admission-to-care <30 min; delayed care = admission-to-care >1 h. “Any pathway delay” = presence of decision delay, reaching delay, transport-access problem, or uncoordinated referral. IQR = interquartile range.

**Table 2.** Admission-to-care time by severe adverse outcome (N = 169)

| Time to neonatal care | Severe (n = 136) | Not severe (n = 33) | $\chi^2$ | p |
| --- | --- | --- | --- | --- |
| <30 minutes | 90 (80.4%) | 22 (19.6%) | 3.95 | 0.139 |
| 30–60 minutes | 35 (87.5%) | 5 (12.5%) |  |  |
| >1 hour | 11 (64.7%) | 6 (35.3%) |  |  |

Note. Values are n (%) within each time category. $\chi^2$ = Pearson chi-square with 2 degrees of freedom, computed across the full 3 × 2 table.

**Table 3.** Association of care-pathway delay indicators with severe adverse outcome, by delay stage (N = 169)

| Delay indicator (present) | Indicator present: severe/total (%) | Indicator absent: severe/total (%) | Crude OR [95% CI] | p |
| --- | --- | --- | --- | --- |
| **Delay 1 — Deciding to seek care** |  |  |  |  |
| Decision-to-seek-care delay (>6 h) | 13/23 (56.5%) | 123/146 (84.2%) | 0.24 [0.10, 0.62] | 0.004 |
| **Delay 2 — Reaching the hospital** |  |  |  |  |
| Transport-access problem | 36/55 (65.5%) | 100/114 (87.7%) | 0.27 [0.12, 0.58] | 0.001 |
| Long distance to facility (>10 km) | 49/58 (84.5%) | 87/111 (78.4%) | 1.50 [0.65, 3.49] | 0.342 |
| Referred / outborn | 72/83 (86.7%) | 64/86 (74.4%) | 2.25 [1.01, 5.00] | 0.043 |
| **Delay 3 — Receiving adequate care** |  |  |  |  |
| Delayed care initiation (>1 h) | 11/17 (64.7%) | 125/152 (82.2%) | 0.40 [0.13, 1.16] | 0.105 |
| Essential drug/feed shortage | 85/99 (85.9%) | 51/70 (72.9%) | 2.26 [1.04, 4.90] | 0.036 |
| **Cross-cutting** |  |  |  |  |
| Any pathway delay present | 51/72 (70.8%) | 85/97 (87.6%) | 0.34 [0.16, 0.76] | 0.006 |

Note. Odds ratios (OR) are crude and unadjusted; 95% CI = 95% confidence interval. p-values are from the Pearson chi-square test, or the Fisher exact test where an expected cell count was <5 (decision delay and delayed care initiation). Inverse (OR < 1) associations for decision delay, transport problems, and any pathway delay most plausibly reflect confounding by indication and reverse causation (the sickest neonates were prioritised for rapid transfer and care) and should not be interpreted as protective effects of delay. These are descriptive associations, not an adjusted determinants model.

**Table 4.** Qualitative themes from key-informant interviews (n = 9), organised by delay.

| Delay | Theme and subthemes | Representative provider account |
| --- | --- | --- |
| **Delay 1** | Caregiver danger-sign recognition and household decision delay (recognition of newborn danger signs; household/partner decision-making; maternal autonomy; cultural beliefs) | *“A mother may say the baby was just ‘sleeping too much’ or ‘not breastfeeding well,’ but to us that is already serious.” (Nurse, 5 years’ experience, KII 02)*<br>*“When the mother is young or dependent on family members, the decision to come to hospital may not be immediate.” (Paediatrician, KII 01)* |
| **Delay 2** | Transport, distance, and referral coordination (cost and night-time travel; long distances; incomplete referral notes; absent pre-arrival calls; poor pre-referral stabilisation) | *“Some babies are referred on motorbikes or public vehicles, wrapped in ordinary clothes, with no oxygen or warmth. By the time they arrive, the situation is already worse.” (Paediatrician, KII 01)*<br>*“Sometimes the referral note only says ‘premature baby’ without temperature, blood sugar, treatment given, or Apgar score. This makes us repeat assessment and lose time.” (Nurse, 5 years’ experience, KII 02)* |
| **Delay 3** | Triage and first-hour stabilisation; facility readiness (first-hour care and triage under congestion; staffing and workload; warm chain and incubators; infection prevention; drug and equipment supply; monitoring and documentation) | *“Newborn care should be treated as an emergency service, because for these small babies, even one hour of delay can change the outcome.” (Paediatrician, KII 01)*<br>*“Staff can do their best, but without enough personnel, equipment, and referral coordination, some preventable complications will continue.” (Newborn-unit manager, KII 09)* |

*Note.* Derived from thematic analysis of nine key-informant interviews (one paediatrician, five nurses, two clinical officers, one newborn-unit manager; 2–10 years’ neonatal experience). Representative quotations are verbatim extracts from the key-informant transcripts, attributed by cadre and interview number; the full coded transcripts are held in the study’s qualitative audit trail. KII = key-informant interview.

**Table 5.** Delay-structured mixed-methods joint matrix.

| Delay | Quantitative indicator (value; association) | Qualitative theme (KII) | Integrated interpretation |
| --- | --- | --- | --- |
| **Delay 1** | Decision delay >6 h: 13.6% (OR 0.24, $p = 0.004$, inverse); mother sole decider: 47.3% | Danger-sign recognition gaps; “just sleepy”; household decision-making; cultural beliefs | The pathway begins at home; recognition and empowerment gaps are real, but the measured delay is confounded because the sickest are brought fastest. |
| **Delay 2** | Referred/outborn: 49.1% (OR 2.25, $p = 0.043$); transport problem: 32.5%; distance >10 km: 34.3%; referral without note: 26.5% | Transport/distance barriers; cold arrivals; incomplete notes; no pre-arrival calls; weak pre-referral stabilisation | Convergent for referral: outborn status marks accumulated vulnerability and a coordination failure. Transport-problem direction is confounded by indication. |
| **Delay 3** | Prompt care <30 min: 66.3% (time not associated, $p = 0.139$); drug/feed shortage: 58.6% (OR 2.26, $p = 0.036$); warm chain: 92.9%; NBU admission: 94.7% | First-hour and triage criticality; staffing/workload; warm chain and incubators; supply gaps; monitoring/documentation | Timeliness at the facility is relatively strong, but supply readiness raises risk; NBU admission largely reflects severity, not poor care. |

*Note.* OR = crude odds ratio for severe adverse outcome; KII = key-informant interview; NBU = newborn unit. Directions marked “inverse/confounded” are discussed in the text as reflecting confounding by indication and reverse causation.

### Delay 1: deciding to seek care

Care was sought immediately by 47.3% of families and within 6 h by a further 39.1%, while 13.6% decided only after more than 6 h (the Delay 1 indicator). The final decision to seek care was made by the mother alone in 47.3% of cases, jointly in 28.4%, by a partner in 15.4%, and by another person in 8.9%; the mother was involved in the decision in 75.7%. Decision-to-seek-care delay was uncommon relative to downstream barriers, but—as the qualitative strand makes clear—it marks the point at which the pathway begins at home. Providers traced this less to indifference than to household dynamics and belief: “Some families believe a small baby can be managed at home with warmth. Others first consult relatives. In some homes, the mother cannot decide alone.” (Nurse, 10 years’ experience, KII 06).

### Delay 2: reaching the hospital

Barriers to reaching care were the most frequent in the cohort. Most families lived rurally (58.6%), and 34.3% were more than 10 km from a facility. Transport was predominantly by motorbike (39.6%) or private car (37.3%), with public transport (18.9%), travel on foot (2.4%), and ambulance (1.8%) far less common; a transport-access problem was reported by 32.5%. Although actual travel time exceeded 2 h in only 1.2% (the Delay 2 travel indicator), nearly half of all neonates (49.1%) were referred/outborn, and among these referred neonates 26.5% (22/83) arrived without a referral note—an indicator of uncoordinated referral. Delay 2 thus combined structural access barriers with a substantial, poorly coordinated referral load. Providers described how the journey itself compounded risk: “If the baby is referred and has been on the road for long, they often arrive with several problems at once: cold, hypoglycemic, and breathing poorly.” (Clinical officer, KII 08).

### Delay 3: receiving adequate care after arrival

After arrival, the facility initiated neonatal care within 30 minutes for 66.3% of neonates, within 30–60 minutes for 23.7%, and after more than 1 h for 10.1% (the Delay 3 indicator). Nearly all neonates were admitted to the newborn unit (94.7%) and had skilled personnel available at admission (94.7%), and the warm chain was maintained in 92.9%. The most prevalent facility-readiness gap was a shortage of essential neonatal drugs or feeds, affecting 58.6% of admissions. Timeliness at the facility was therefore relatively strong, but supply readiness was not. Providers attributed the residual delays more to logistics than to clinical hesitation: “Admission procedures, getting a bed space, finding equipment, and starting treatment can take longer than desired.” (Nurse, 10 years’ experience, KII 06).

### Delay indicators and the severe outcome

The proportion of severe outcomes did not differ significantly across categories of admission-to-care time (Table 2; χ²(2) = 3.95, p = 0.139), with severe outcomes common even among neonates seen within 30 minutes (80.4%). When individual delay indicators were related to the severe outcome (Table 3), a consistent and important pattern emerged. Referral/outborn status was associated with higher odds of a severe outcome (crude OR 2.25, 95% CI 1.01–5.00; p = 0.043), as was a shortage of essential drugs or feeds (OR 2.26, 95% CI 1.04–4.90; p = 0.036). In contrast, decision-to-seek-care delay (OR 0.24; p = 0.004), transport-access problems (OR 0.27; p = 0.001), and the presence of any pathway delay (OR 0.34; p = 0.006) were each associated with a lower proportion of severe outcomes.

These inverse associations are counter-intuitive and should not be read as protective effects of delay. They most plausibly reflect confounding by indication and reverse causation: the smallest and most obviously compromised neonates were recognised early, prioritised for rapid transfer, and fast-tracked into care, so that the most severe cases are concentrated among the shortest measured delays. The two indicators that behaved in the expected direction—referral status and supply shortage—are precisely those that capture accumulated vulnerability and facility-readiness rather than the speed of an individual family’s journey. This interpretive caution is carried through the joint display (Table 5) and the discussion.

### Provider perspectives on the care pathway

Provider accounts, coded directly to the three delays, explained how the pathway is experienced at the bedside (Table 4). For Delay 1, informants described caregivers delaying because early danger signs in a small neonate were subtle or misread—“a small baby thought to be just sleepy”— and because the final decision to seek care often rested with a partner or elder; some families also held beliefs that a very small or sick baby was a bad omen. For Delay 2, providers repeatedly linked transport cost, night-time travel, and long distances to neonates “arriving cold,” and singled out referral coordination—incomplete notes, absent pre-arrival calls, and inadequate pre-referral stabilisation—as a driver of cold, unstable, and poorly documented admissions. For Delay 3, informants emphasised the criticality of the first hour and of triage under congestion, and described staffing and workload pressure (high nurse-to-baby ratios, night and weekend burnout), dependence on kangaroo mother care where incubators were insufficient, infection-prevention challenges in a crowded unit, intermittent shortages of drugs and equipment (oxygen, CPAP, feeds, antibiotics), and monitoring and documentation that were sometimes sacrificed during emergencies.

### Integrated delay matrix

The delay-structured joint display (Table 5) reads the quantitative indicators against the corresponding provider themes within each delay. The two strands converge most clearly on referral: nearly half the cohort was outborn, referral was statistically associated with a severe outcome, and providers described referral coordination as a recurrent point of failure. They also converge on facility-readiness gaps, where supply shortages were both measured and described. Where the strands diverge—decision delay and transport problems appearing statistically ‘protective’ while providers described them as harmful—the divergence is itself informative, pointing to confounding by indication rather than to a true benefit of delay. Finally, several strong qualitative themes (triage under congestion, staffing adequacy, monitoring, infection prevention, admission temperature) had no matched quantitative variable, marking clear measurement gaps for future data collection.

## Discussion

Applying the Three Delays Model [8,9] to LBW neonates at a Kenyan county referral hospital, we found that the pathway’s barriers were concentrated before arrival. Delay 2—reaching the hospital—carried the most prevalent barriers (long distance in 34.3%, transport problems in 32.5%, and referral/outborn status in 49.1%), while the facility itself initiated care promptly (<30 min) in two-thirds of neonates. The single delay indicator that behaved as a marker of risk in the expected direction was referral/outborn status (OR 2.25), together with facility-readiness measured as essential drug or feed shortage (OR 2.26). Provider narratives, coded to the same three delays, converged on referral coordination and first-hour stabilisation as the recurrent points of failure. This ordering—barriers massed in the second delay while the receiving facility responded relatively quickly—mirrors newborn applications of the model elsewhere in sub-Saharan Africa, in which reaching care and being stabilised for it, rather than the family’s initial decision, dominate the pathway to neonatal death [9–12].

A central argument follows from these data: a referred LBW neonate represents more than a transport statistic. Half of the cohort was outborn, a quarter of referred neonates arrived without a referral note, and referral status was associated with a higher likelihood of a severe outcome. Read alongside the provider accounts of cold, unstable, and poorly documented arrivals, referral status is best understood as a summary marker of accumulated vulnerability: it bundles delayed decision-making at home, transport constraints, incomplete pre-referral stabilisation, and a facility-response gap into a single point in the pathway [9,15]. This reading is consistent with the original logic of the model, in which survival depends on an unbroken chain of timely action across households, transporters, and facilities [8], a framework whose applicability to access barriers in low- and middle-income settings has been repeatedly validated and critically refined [14]. It also accords with assessments of Kenyan neonatal referral showing that transport is frequently undertaken without thermal protection, oxygen, or structured communication, so that infants arrive already compromised [15], and with the wider observation that neonates in low- and middle-income countries remain “too small to be seen”, their outcomes shaped as much by where and how they are moved as by their intrinsic biology [7]. Strengthening the referral transition—advance communication, pre-referral warmth and glucose, and structured handover—may therefore yield more than interventions aimed at any single family’s decision speed.

Several delay indicators—decision delay, transport problems, and the composite ‘any delay’— were statistically associated with fewer severe outcomes. Taken at face value this is implausible, and we interpret it as confounding by indication and reverse causation rather than as a protective effect of delay. In a referral setting, the most obviously compromised neonates are recognised early and moved and treated fastest, concentrating severe outcomes among the shortest measured intervals; conversely, some neonates who tolerated a longer journey may have been comparatively more stable at the outset. The same interpretive caution applies to newborn-unit admission, which reflects illness severity rather than poor care, and it is why we deliberately kept the analysis descriptive and did not construct an adjusted delay model. The lesson is methodological as much as clinical: in facility-based neonatal pathway studies, simple delay–outcome associations can invert, and they must be read through the lens of triage behaviour and the mixed-methods joint display rather than in isolation. Comparable applications of the model have similarly cautioned that the delay which is easiest to measure is not necessarily the one most responsible for death, and that facility, verbal, and social-autopsy data require careful causal interpretation [11,13].

For this setting, the most actionable break points sit at the Delay 2–Delay 3 interface: the referral transition and the first hour after arrival. Timeliness at the facility was already relatively strong, so the marginal gains lie less in speeding admission and more in what happens around it—reliable pre-referral stabilisation and communication, and a dependable warm-chain, glucose, oxygen, and antibiotic response in the first hour, supported by consistent essential supplies [4,16,20]. Delay 1 remains important as the origin of the pathway and is addressable through community danger-sign recognition and caregiver empowerment, but on these data it is neither the most prevalent nor, once confounding is accounted for, the clearest quantitative signal. Framing the question as which delay matters most locally, rather than treating the three as interchangeable, follows directly from newborn applications of the model that ask whether prevailing programmes are in fact focused on the delays that drive mortality in a given place [11].

These findings are consistent with a body of work applying delay frameworks to newborn survival in sub-Saharan Africa and South Asia, in which pre-hospital decision and transport barriers and weak referral systems feature prominently [9–13], and with facility-based assessments in Kenya documenting gaps in inpatient newborn care capacity, supplies, and nursing workload [17–19]. The prominence of hypothermia on arrival [5], the dependence on kangaroo mother care where incubators are limited [21–23], and the risk associated with supply shortages [24] all echo regional evidence on small and sick newborn care [4]. Hypothermia among Kenyan newborn-unit admissions has specifically been shown to cluster in low-birth-weight infants and to predict inpatient mortality, reinforcing why the warm chain recurred so strongly in provider accounts [25]. What our mixed-methods design adds is the explicit, delay-structured integration that both corroborates the referral and supply findings and exposes why crude delay associations can mislead.

The results point to concrete, pathway-specific actions. For Delay 1, community and caregiver education on newborn danger signs and support for prompt, autonomous care-seeking. For Delay 2, strengthened referral communication through advance notification and complete referral notes, pre-referral stabilisation—especially thermal care and glucose—and more reliable emergency transport [15]. For Delay 3, rapid, protocol-driven triage; a dependable first-hour bundle of warm-chain maintenance, glucose and oxygen assessment, and timely antibiotics [16,20]; and consistent availability of essential neonatal drugs, feeds, and equipment, supported by monitoring and documentation that hold up under emergency workload [19]. These measures are neither novel nor costly; the recurring lesson from low- and middle-income settings is that outcomes improve when established interventions for small and sick newborns are delivered reliably rather than intermittently [4,26]. Because they are pathway and system measures, they complement—rather than duplicate—the biological risk-stratification and nursing-practice questions addressed elsewhere in this body of work.

Several strengths and limitations frame these conclusions. Strengths include the explicit theoretical structure, the mixed-methods design with a delay-structured joint display, verification of all quantitative indicators against the primary dataset, and transparent handling of confounded associations. The design is nonetheless cross-sectional and single-site, so temporal and causal inferences are not warranted and generalisability is limited. The severe outcome was highly prevalent (80.5%), leaving a small unaffected group (n = 33) and wide confidence intervals; association testing was therefore kept descriptive and unadjusted by design. Some clinically salient pathway constructs—admission temperature, danger-sign recognition, triage time under congestion, staffing adequacy, and monitoring intensity—were not captured as quantitative variables and were available only qualitatively, a measurement gap that future pathway studies should close. Finally, several delay indicators are self-reported and subject to recall, and the observed inverse associations underscore that confounding by indication is a structural feature of facility-based delay data that must be addressed in any subsequent analytic modelling [11].

## Conclusions

Among low-birth-weight neonates at a Kenyan county referral hospital, care-pathway barriers accumulated before arrival, and referral/outborn status—more than any single measure of a family’s decision speed—marked accumulated vulnerability and a coordination failure associated with severe outcomes. Facility timeliness was relatively strong, but supply readiness and the first-hour response were not. Reducing severe adverse outcomes for the smallest neonates therefore requires shortening specific, identifiable delays across the full pathway—above all strengthening the referral transition from lower facilities and safeguarding the fragile first hour after arrival— rather than reproducing a full determinants model.

## Additional Information

### Consent for publication

Not applicable; no individually identifiable data are presented.

### Data availability

The de-identified dataset and analysis syntax are available from the corresponding author on reasonable request, subject to ethical-approval conditions. A delay-indicator codebook is provided as Supporting Information (S1).

## Authors’ contributions

JC conceived and conducted the study and led data collection. MSS supervised the design and analysis and drafted the manuscript. VKM and EM contributed to data curation, interpretation, and critical revision. All authors read and approved the final manuscript.

## Data Availability

Data is accessible through OSF platform:https://doi.org/10.17605/OSF.IO/NMKAU

## Acknowledgements

The authors thank the mothers and providers who participated, and the management and staff of the Kericho County Referral Hospital newborn unit.

## Abbreviations

ANC: antenatal care
CI: confidence interval
COREQ: Consolidated Criteria for Reporting Qualitative Research
CPAP: continuous positive airway pressure
GRAMMS: Good Reporting of a Mixed Methods Study
IQR: interquartile range
KCRH: Kericho County Referral Hospital
KII: key-informant interview
KMC: kangaroo mother care
LBW: low birth weight
NBU: newborn unit
NICU: neonatal intensive care unit
OR: odds ratio
STROBE: Strengthening the Reporting of Observational Studies in Epidemiology

## Supporting information

S1 Codebook. Delay-indicator codebook (indicator, source variable, derivation, and denominator) for all indicators reported in Table 1.

## References

1. Blencowe H, Krasevec J, de Onis M, Black RE, An X, Stevens GA, et al. National, regional, and worldwide estimates of low birthweight in 2015. Lancet Glob Health. 2019;7(7):e849–e860.

2. Lawn JE, Blencowe H, Oza S, You D, Lee ACC, Waiswa P, et al. Every Newborn: progress, priorities, and potential beyond survival. Lancet. 2014;384(9938):189–205.

3. Hug L, Alexander M, You D, Alkema L; UN Inter-agency Group for Child Mortality Estimation. National, regional, and global levels and trends in neonatal mortality between 1990 and 2017. Lancet Glob Health. 2019;7(6):e710–e720.

4. World Health Organization. Survive and thrive: transforming care for every small and sick newborn. Geneva: World Health Organization; 2019.

5. Lunze K, Bloom DE, Jamison DT, Hamer DH. The global burden of neonatal hypothermia: systematic review of a major challenge for newborn survival. BMC Med. 2013;11:24.

6. Seale AC, Blencowe H, Manu AA, Nair H, Bahl R, Qazi SA, et al. Estimates of possible severe bacterial infection in neonates in sub-Saharan Africa, south Asia, and Latin America. Lancet Infect Dis. 2014;14(8):731–741.

7. Rosa-Mangeret F, Benski AC, Golaz A, Zala PZ, Kyokan M, Wagner N, et al. 2.5 million annual deaths—are neonates in low- and middle-income countries too small to be seen? A bottom-up overview on neonatal morbi-mortality. Trop Med Infect Dis. 2022;7(5):64.

8. Thaddeus S, Maine D. Too far to walk: maternal mortality in context. Soc Sci Med. 1994;38(8):1091– 1110.

9. Waiswa P, Kallander K, Peterson S, Tomson G, Pariyo GW. Using the three delays model to understand why newborn babies die in eastern Uganda. Trop Med Int Health. 2010;15(8):964–972.

10. Upadhyay RP, Rai SK, Krishnan A. Using three delays model to understand the social factors responsible for neonatal deaths in rural Haryana, India. J Trop Pediatr. 2013;59(2):100–105.

11. Kaselitz EB, Cunningham-Rhoads B, Aborigo RA, Williams JEO, James KH, Moyer CA. Neonatal mortality in rural northern Ghana and the three delays model: are we focusing on the right delays? Trop Med Int Health. 2021;26(5):582–590.

12. Salmen CR, Ndunyu L, Ssenkusu JM, Marshall D, DesLauriers N, Anebarassou AV, et al. Falling through the net: an adaptive assessment of the ‘Three Delays’ encountered by patients seeking emergency maternal and neonatal care within a remote health system on Lake Victoria, Kenya. Glob Public Health. 2022;17(9):2156–2175.

13. Wilmot E, Yotebieng M, Norris A, Ngabo F. Missed opportunities in neonatal deaths in Rwanda: applying the three delays model in a cross-sectional analysis of neonatal death. Matern Child Health J. 2017;21(5):1121–1129.

14. Actis Danna V, Bedwell C, Wakasiaka S, Lavender T. Utility of the three-delays model and its potential for supporting a solution-based approach to accessing intrapartum care in low- and middle-income countries. A qualitative evidence synthesis. Glob Health Action. 2020;13(1):1819052.

15. Wainaina J, Irimu G, English M, Mbaire E, Waiyego M, Manyasi C, et al. Assessment of neonatal referral infrastructure and clinical characteristics of referred neonates in three first referral hospitals in Nairobi County, Kenya. Wellcome Open Res. 2023;8:126.

16. Ray S, Sundaram V, Dutta S, Kumar P. Ensuring administration of first dose of antibiotics within the golden hour of management in neonates with sepsis. BMJ Open Qual. 2021;10(Suppl 1):e001365.

17. Aluvaala J, Nyamai R, Were F, Wasunna A, Kosgei R, Karumbi J, et al. Assessment of neonatal care in clinical training facilities in Kenya. Arch Dis Child. 2015;100(1):42–47.

18. Murphy GAV, Gathara D, Abuya N, Mwachiro J, Ochola S, Snow R, et al. What capacity exists to provide essential inpatient care to small and sick newborns in a high mortality urban setting? A cross-sectional study in Nairobi City County, Kenya. PLoS One. 2018;13(4):e0196585.

19. Gathara D, Serem G, Murphy GAV, Obengo A, Tallam E, Jackson D, et al. Missed nursing care in newborn units: a cross-sectional direct observational study. BMJ Qual Saf. 2020;29(1):19–30.

20. Enweronu-Laryea C, Dickson KE, Moxon SG, Simen-Kapeu A, Nyange C, Niermeyer S, et al. Basic newborn care and neonatal resuscitation: a multi-country analysis of health system bottlenecks and potential solutions. BMC Pregnancy Childbirth. 2015;15(Suppl 2):S4.

21. Lawn JE, Mwansa-Kambafwile J, Horta BL, Barros FC, Cousens S. Kangaroo mother care to prevent neonatal deaths due to preterm birth complications. Int J Epidemiol. 2010;39(Suppl 1):i144–i154.

22. Boundy EO, Dastjerdi R, Spiegelman D, Fawzi WW, Missmer SA, Lieberman E, et al. Kangaroo mother care and neonatal outcomes: a meta-analysis. Pediatrics. 2016;137(1):e20152238.

23. WHO Immediate KMC Study Group. Immediate “kangaroo mother care” and survival of infants with low birth weight. N Engl J Med. 2021;384(21):2028–2038.

24. Moxon SG, Lawn JE, Dickson KE, Simen-Kapeu A, Gupta G, Deorari A, et al. Inpatient care of small and sick newborns: a multi-country analysis of health system bottlenecks and potential solutions. BMC Pregnancy Childbirth. 2015;15(Suppl 2):S7.

25. Wainaina J, Ogero M, Mumelo L, Wairoto K, Mbevi G, Tuti T, et al. Hypothermia amongst neonatal admissions in Kenya: a retrospective cohort study assessing prevalence, trends, associated factors, and its relationship with all-cause neonatal mortality. Front Pediatr. 2024;12:1272104.

26. Kleinhout MY, Stevens MM, Osman KA, Adu-Bonsaffoh K, Groenendaal F, Biza Zepro N, et al. Evidence-based interventions to reduce mortality among preterm and low-birthweight neonates in low-income and middle-income countries: a systematic review and meta-analysis. BMJ Glob Health. 2021;6(2):e003618.

27. Blencowe H, Cousens S, Oestergaard MZ, Chou D, Moller AB, Narwal R, et al. National, regional, and worldwide estimates of preterm birth rates in the year 2010 with time trends. Lancet. 2012;379(9832):2162–2172.

28. Chawanpaiboon S, Vogel JP, Moller AB, Lumbiganon P, Petzold M, Hogan D, et al. Global, regional, and national estimates of levels of preterm birth in 2014. Lancet Glob Health. 2019;7(1):e37–e46.

29. World Health Organization. Standards for improving quality of maternal and newborn care in health facilities. Geneva: World Health Organization; 2016.

30. Fleischmann-Struzek C, Goldfarb DM, Schlattmann P, Schlapbach LJ, Reinhart K, Kissoon N. The global burden of paediatric and neonatal sepsis: a systematic review. Lancet Respir Med. 2018;6(3):223–230.

31. Mgawadere F, Unkels R, Kazembe A, van den Broek N. Factors associated with maternal mortality in Malawi: application of the three delays model. BMC Pregnancy Childbirth. 2017;17(1):219.

32. Combs Thorsen V, Sundby J, Malata A. Piecing together the maternal death puzzle through narratives: the three delays model revisited. PLoS One. 2012;7(12):e52090.

33. Kalter HD, Salgado R, Babille M, Koffi AK, Black RE. Social autopsy for maternal and child deaths: a comprehensive literature review to examine the concept and the development of the method. Popul Health Metr. 2011;9:45.

34. Say L, Chou D, Gemmill A, Tunçalp Ö, Moller AB, Daniels J, et al. Global causes of maternal death: a WHO systematic analysis. Lancet Glob Health. 2014;2(6):e323–e333.

35. Tunçalp Ö, Were WM, MacLennan C, Oladapo OT, Gülmezoglu AM, Bahl R, et al. Quality of care for pregnant women and newborns—the WHO vision. BJOG. 2015;122(8):1045–1049.

36. Kenya National Bureau of Statistics, ICF. Kenya Demographic and Health Survey 2022. Nairobi and Rockville (MD): KNBS and ICF; 2023.

37. Ministry of Health, Republic of Kenya. Basic Paediatric Protocols. 5th ed. Nairobi: Ministry of Health; 2022.

38. von Elm E, Altman DG, Egger M, Pocock SJ, Gøtzsche PC, Vandenbroucke JP; STROBE Initiative. The Strengthening the Reporting of Observational Studies in Epidemiology (STROBE) statement: guidelines for reporting observational studies. Lancet. 2007;370(9596):1453–1457.

39. Tong A, Sainsbury P, Craig J. Consolidated criteria for reporting qualitative research (COREQ): a 32-item checklist for interviews and focus groups. Int J Qual Health Care. 2007;19(6):349–357.

40. O’Cathain A, Murphy E, Nicholl J. The quality of mixed methods studies in health services research. J Health Serv Res Policy. 2008;13(2):92–98.

41. Braun V, Clarke V. Using thematic analysis in psychology. Qual Res Psychol. 2006;3(2):77–101.

